# Correlation between prevalence of tobacco smoking and risk and severity of COVID-19 at the national level in the European Union: an ecological study

**DOI:** 10.1101/2020.04.28.20083352

**Authors:** Adrián González-Marrón, Jose M Martínez-Sánchez

## Abstract

The Angiotensin-converting enzyme 2 (ACE2) protein is the receptor for different coronaviruses, including Severe acute respiratory syndrome coronavirus 2 (SARS-CoV-2), the causative agent of Coronavirus disease 2019 (COVID-19). Previous studies suggested the hypothesis that nicotine could downregulate the expression of the ACE2. Due to the high level of nicotine intake, the objective of this preliminary study was to assess, at the ecological level, the correlation between tobacco smoking and the attack rate and severity of COVID-19 in the European Union (EU). We have found that there is a statistically significant negative correlation between the age-standardized prevalence of tobacco smoking and the attack rate of COVID-19 in member states of the EU [Spearman’s correlation coefficient = −0.476 (95% confidence interval −0.117, −0.725) (p-value = 0.012)], meaning that in member states with a higher age-standardized prevalence of tobacco smoking the attack rate of COVID-19 has been so far lower. Further research is needed to understand the possible effect of nicotine exposure in the expression of the ACE2 protein.

## Introduction

Severe acute respiratory syndrome coronavirus 2 (SARS-CoV-2) is a newly discovered virus, belonging to the family *Coronaviridae* (Gorbalenya *et al*., 2020), and causative agent of the disease Coronavirus disease 2019 (COVID-19). As of April 14^th^, around 2 million cases and 120,000 deaths of COVID-19 have been confirmed globally (Johns Hopkins University & Medicine, 2020). Although the origin of the virus was the city of Wuhan, China, the center of the pandemic of COVID-19 is currently in Europe, with four out of the five countries accumulating more deaths worldwide located in this continent (i.e., Italy, Spain, France and United Kingdom).

The Angiotensin-converting enzyme 2 (ACE2) protein is the receptor for different coronaviruses, including SARS-CoV-2 (Hoffmann *et al*., 2020). However, evidence is still sparse on the effect of different exposures on the interaction ACE2 – SARS-CoV-2. Specifically, there is not conclusive evidence on the regulation of the ACE2 in the lungs after exposure to nicotine (Oakes *et al*., 2018) (Brake *et al*., 2020), one of the thousands of substances present in the tobacco smoke (Rodgman and Perfetti, 2013), and how this regulation could impact the outcomes associated with the COVID-19 infection, eventually. Preliminary epidemiological data indicate that the prevalence of current smokers in hospitalized patients in China due to COVID-19 was lower than at the national level (Farsalinos *et al*., 2020) and that current smokers with COVID-19 seem to be at higher risk of negative progression than non-smokers (Vardavas and Nikitara, 2020), although other authors concluded that severity is not associated with active smoking (Lippi and Henry, 2020).

Assuming the hypothesis that smokers, due to the high level of nicotine intake, underexpress the ACE2 protein, countries with a higher prevalence of smoking may have lower attack rates of COVID-19. Hence, the objective of this preliminary study was to assess the correlation between tobacco smoking and the attack rate and severity of COVID-19 at the national level in the European Union (EU).

## Material and methods

This is an ecological study. The unit of analysis is each of the 27 member states of the EU. Data on confirmed cases and deaths due to the COVID-19 for each member state of the EU were retrieved from the Johns Hopkins University Coronavirus Resource Center (Dong *et al*., 2020) using metadata accessible from GitHub until April 14^th^ 2020 (Johns Hopkins University & Medicine, 2020). Age-standardized prevalence of tobacco smoking at the national level among persons 15 years and older in 2016 were retrieved from the World Health Statistics on Tobacco control (World Health Organization, 2018). Population on January 1^st^ 2019 for each of the member states of the EU was retrieved from Eurostat (Eurostat, 2019).

The attack rate (cumulative cases/100,000 population) and the case fatality ratio (CFR) (deaths/cumulative cases) were computed for each member state. Due to the non-normality of data, we computed Spearman’s rank correlation coefficients, and their 95% confidence intervals (95% CI), of the age-standardized prevalence of tobacco smoking and the attack rate and of the age-standardized prevalence of tobacco smoking and the CFR for each of the member states. Univariate linear regression models were computed to obtain crude regression coefficients (β), and their 95% CI, for both associations. Scatterplots were designed to graphically depict both associations. P-values were considered significant when <0.05.

## Results

Table 1 contains descriptive data at the national level. The Spearman’s rank correlation coefficient of the age-standardized prevalence of tobacco smoking and the attack rate was −0.476 (95% CI −0.117, −0.725) (p-value = 0.012), with β = −7.325 (95% CI −15.028, 0.378) (p-value = 0.061) (Figure 1, panel A). The Spearman’s rank correlation coefficient between the age-standardized prevalence of tobacco smoking and the CFR was −0.205 (95% CI −0.543, 0.196) (p-value = 0.304), with β = −0.119 (95% CI −0.377, 0.139) (p-value = 0.351) (Figure 1, panel B).

**Table 1.** Descriptive data at the national level of the member states of the EU for the COVID-19 outbreak as of April 14^th^ 2020.

|  | Deaths | Cases | Population<br>(2019) | Age-standardized<br>prevalence of tobacco<br>smoking | Attack rate (per<br>100,000<br>population) | Case<br>fatality<br>rate (%) |
| --- | --- | --- | --- | --- | --- | --- |
| Austria | 384 | 14,226 | 8,858,775 | 29.7 | 160.59 | 2.70 |
| Belgium | 4,157 | 31,119 | 11,467,923 | 28.3 | 271.36 | 13.36 |
| Bulgaria | 35 | 713 | 7,000,039 | 37.3 | 10.19 | 4.91 |
| Croatia | 31 | 1,704 | 4,076,246 | 37.1 | 41.80 | 1.82 |
| Cyprus | 12 | 695 | 875,898 | 36.2 | 79.35 | 1.73 |
| Czechia | 161 | 6,111 | 10,649,800 | 34.4 | 57.38 | 2.63 |
| Denmark | 299 | 6,511 | 5,806,081 | 19.1 | 112.14 | 4.59 |
| Estonia | 31 | 1,373 | 1,324,820 | 31.9 | 103.64 | 2.26 |
| Finland | 64 | 3,161 | 5,517,919 | 20.5 | 57.29 | 2.02 |
| France | 15,729 | 130,253 | 67,028,048 | 32.9 | 194.33 | 12.08 |
| Germany | 3,294 | 131,359 | 83,019,214 | 30.7 | 158.23 | 2.51 |
| Greece | 101 | 2,170 | 10,722,287 | 43.7 | 20.24 | 4.65 |
| Hungary | 122 | 1,512 | 9,797,561 | 30.8 | 15.43 | 8.07 |
| Ireland | 406 | 11,479 | 4,904,226 | 24.4 | 234.06 | 3.54 |
| Italy | 21,067 | 162,488 | 60,359,546 | 23.8 | 269.20 | 12.97 |
| Latvia | 5 | 657 | 1,919,968 | 38.3 | 34.22 | 0.76 |
| Lithuania | 29 | 1,070 | 2,794,184 | 29.7 | 38.29 | 2.71 |
| Luxembourg | 67 | 3,307 | 613,894 | 23.5 | 538.69 | 2.03 |
| Malta | 3 | 393 | 493,559 | 25.6 | 79.63 | 0.76 |
| Netherlands | 2,945 | 27,419 | 17,282,163 | 25.9 | 158.65 | 10.74 |
| Poland | 263 | 7,202 | 37,972,812 | 28.2 | 18.97 | 3.65 |
| Portugal | 567 | 17,448 | 10,276,617 | 23.2 | 169.78 | 3.25 |
| Romania | 351 | 6,879 | 19,401,658 | 30.0 | 35.46 | 5.10 |
| Slovakia | 2 | 835 | 5,450,421 | 30.4 | 15.32 | 0.24 |
| Slovenia | 56 | 1,220 | 2,080,908 | 22.6 | 58.63 | 4.59 |
| Spain | 18,056 | 172,541 | 46,934,632 | 29.4 | 367.62 | 10.46 |
| Sweden | 1,033 | 11,445 | 10,230,185 | 18.9 | 111.87 | 9.03 |

**Figure 1.**
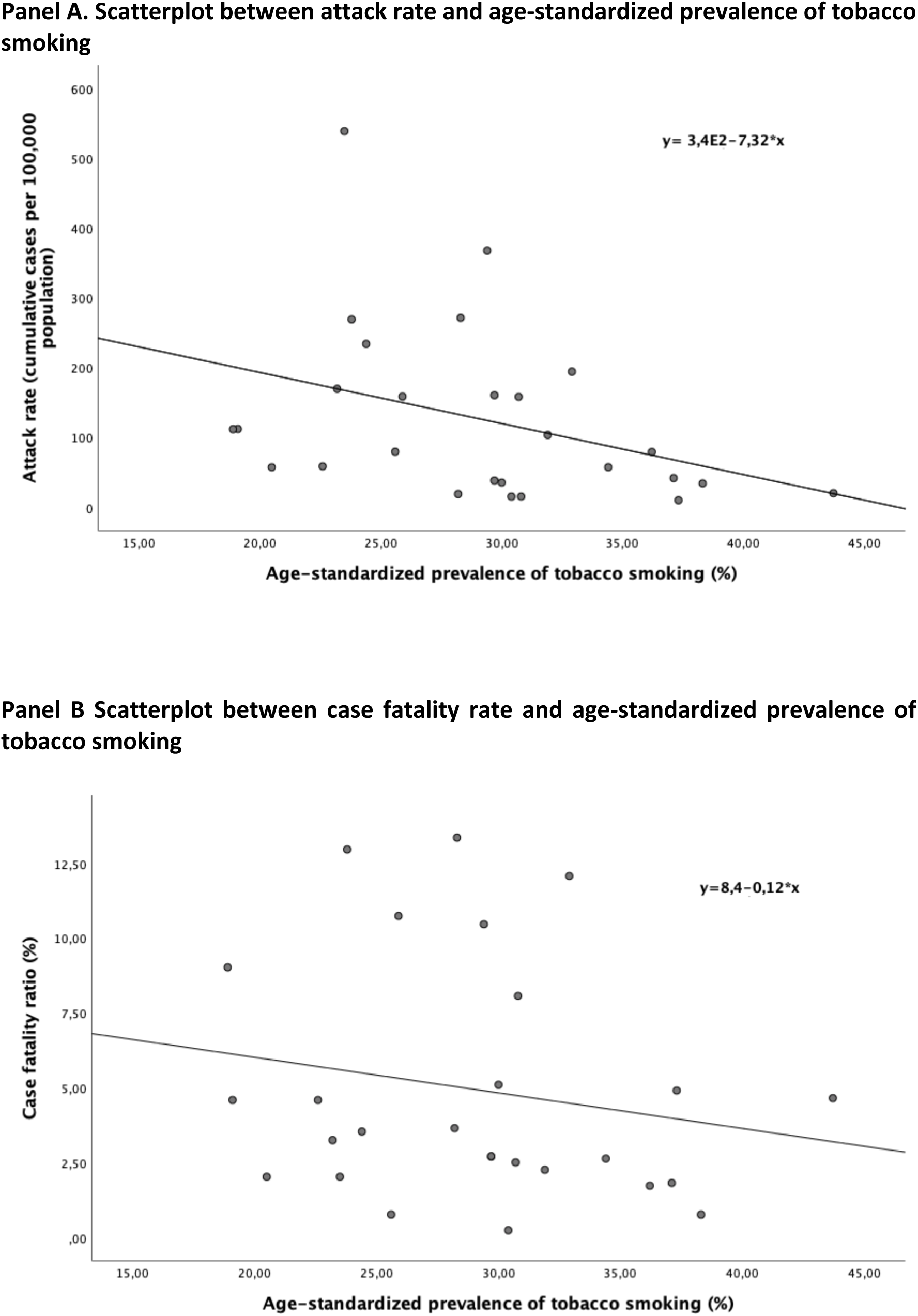
Scatterplots at the national level in the member states of the EU.

## Discussion

Our preliminary analyses show that there is a statistically significant negative correlation between the age-standardized prevalence of tobacco smoking and the attack rate of COVID-19 at the national level in the EU, meaning that in member states with a higher age-standardized prevalence of tobacco smoking the attack rate of COVID-19 has been so far lower. These preliminary results would support the hypotheses, at the ecological level only, that nicotine could downregulate the expression of ACE2 (Oakes *et al*., 2018). Importantly, due to the ongoing nature of this recent pandemic and the shortness of available data, our results should be taken with caution.

We want to highlight that tobacco smoke contains thousands of substances, not only nicotine, including systemic irritants and toxins, such as hydrocyanic acid, sulfur dioxide, carbon monoxide, ammonia and formaldehyde, as well as around 70 carcinogenic and mutagenic substances, such as arsenic, chromium, nitrosamines and benzopyrene (Rodgman and Perfetti, 2013). In this sense, and as other authors state (Vardavas and Nikitara, 2020)(Gorini *et al*., 2020), in the absence of more data, tobacco smoking should be considered as associated with poorer prognosis of COVID-19.

This study contains some limitations, which should be addressed. Firstly, this is an ecological study, meaning that conclusions at the individual level should not be drawn. Also, data are still preliminary due to the outbreak being still ongoing and adjustment for confounders was not possible (public health measures such as lockdowns, population density of the member states, etc.). Besides, quality and accuracy of numbers on COVID-19 cases and deaths may vary between member states due to differences in capacities in national surveillance systems and definitions of case and death.

In conclusion, with data available as of April 14^th^ 2020, and at the ecological level, in the member states of the EU with higher rates of smoking, the attack rate of COVID-19 has been lower. In this sense, further research is needed to understand the possible effect of nicotine exposure in the expression of the ACE2 protein, and eventually in the outcomes of COVID-19.

## Data Availability

Manuscript using public data.

## Acknowledgement

We would like to thank Prof. Josep Clotet i Erra for sharing with us his research hypothesis that nicotine could downregulate the expression of ACE2. Without his research idea, we would not have done this paper.

## Notes

### Competing Interest Statement

The authors have declared no competing interest.

### Funding Statement

No funding

